# Predictors of treatment outcomes with Auto-titrating CPAP therapy in adults with Obstructive Sleep Apnea

**DOI:** 10.1101/2021.10.21.21265338

**Authors:** Olabimpe S Fashanu, Stuart F Quan

## Abstract

**Objectives:** To determine factors that predict OSA therapy outcomes with auto-titrating positive airway pressure (APAP).

**Methods:** We sequentially grouped patients from a retrospective cohort based on APAP efficacy (sufficiently vs. insufficiently treated; insufficiently treatment defined as residual AHI of ≥ 5); therapy adherence (compliant vs. non-compliant, non-compliance defined as < 70% usage for ≥ 4 hours/night); and therapy outcomes (optimal vs non-optimal (non-optimal outcomes defined as non-compliant and insufficiently treated). We subsequently compared each group.

**Results:** The insufficiently treated were older (68.4 ± 12.5 vs. 60.4 ± 13.1 years, p < 0.01) and had lower BMI (31.9 ± 6.3 vs.37.9 ± 9.1 kg/m^2^, p < 0.01). They had higher baseline central apnea indices (CAI), longer leaks, higher peak pressures and were less compliant.

The non-compliant were younger (61.1 ± 12.6 vs. 65.5 ± 13.2 years, p = 0.03) and comprised more females (56.1 vs. 43.9%, p = 0.04). The leak duration per usage hour was higher in the non-compliant (13.9 ± 42.1 vs. 1.83 ± 3.53 minutes/hour, p < 0.01). The non-optimally treated had lower BMI, longer leaks and shorter nightly usage.

Following multivariate analysis, the aforementioned variables except age were predictive of higher residual AHI, while age and gender predicted compliance.

**Conclusions:** Various demographic and clinical factors were predictive of treatment efficacy and adherence. Overall, lower BMI, longer leaks and shorter nightly usage predicted poor therapy outcomes.

## Introduction

Continuous positive airway pressure (CPAP) is the gold standard in the treatment of obstructive sleep apnea (OSA); obstructive events are virtually eliminated in compliant patients. Until recently, CPAP therapy was primarily delivered via the fixed pressure mode. However, auto-titrating positive airway pressure (APAP) during which airway pressures are automatically adjusted throughout the sleep period is also a recommended option [1] and has become an increasingly popular mode of CPAP delivery for the treatment of uncomplicated OSA.

CPAP treatment outcomes are often limited by poor treatment adherence. To achieve optimal patient outcomes, CPAP efficacy must be paired with adequate compliance with treatment.

Prior studies on positive airway pressure (PAP) outcomes have either been done using a variety of PAP delivery modes [2] or have not documented that the studies were limited to a particular mode of PAP delivery [3]. Although treatment efficacy with APAP is generally comparable to CPAP, with APAP the machine-specific algorithmic automatic adjustment in airway pressure may sometimes be inadequate to result in a sufficient reduction in the apnea hypopnea index (AHI) [4] [5]. The goal of this study was to determine demographic and clinical factors that may predict therapy outcomes with APAP in patients with OSA. Outcomes of interest were treatment effectiveness as determined by a reduction in AHI and adherence. This knowledge is crucial to developing specific interventions targeted at achieving better treatment outcomes.

## Methods

Data were analyzed from a retrospective cohort of adult patients aged ≥ 18 years treated with APAP at the Brigham and Women’s Faulkner Hospital Sleep Clinic between September 2015 and September 2018. As described previously [6], to be included in the study, patients were required to have been treated with APAP and not fixed pressure CPAP, and to have been followed up in the sleep clinic within 18 months of their CPAP titration polysomnogram. Patients who had comorbid sleep related breathing disorders including central sleep apnea, sleep related hypoxemia or hypoventilation were excluded. Of the 922 charts reviewed, a total of 189 patients initially fulfilled study criteria. However, for uniformity of analyses, only patients who were prescribed APAP devices manufactured by Philips Respironics, Murrysville, PA were included in this study. Therefore, 17 patients using other devices were excluded, resulting in a final cohort comprising 172 patients. Demographic, anthropometric and clinical data including 30-day APAP compliance data were extracted from the electronic medical record on eligible study participants. Therapy effectiveness was defined as residual AHI <5 /h) and treatment adherence was defined as APAP device usage of ≥70% of nights for at least 4 h/night over 30 consecutive nights.

Definition of Leaks: There is unfortunately no universal definition of significant unintentional mask leaks, and the definition varies between CPAP manufacturers. Large leaks are generally considered significant when the machine manufacturer is concerned that the leak will impact the proper function of the machine. Philips Respironics APAP compliance data documented the duration of large leaks per day. The manufacturer defines small and large mask leaks as ≤ 1 and >1 hour of large leak per day respectively [7]. This definition may however, be impractical if not clinically correlated given individual variability in hours of usage of APAP. We adjusted the duration of large leaks per day for both hours of nightly usage and for pressure delivery using the pressure at which the APAP device spent 90% of the time at or below (90th percentile pressure).

### Statistical Analysis

Baseline characteristics of study participants were summarized as means and standard deviations for continuous and interval variables, and as percentages for categorical variables. To evaluate factors impacting therapy effectiveness, patients were divided into 2 groups based on whether their residual AHI was ≥ or < 5 /hour. Similarly, to assess treatment adherence, patients also were assigned into 2 groups based on whether they met the treatment adherence criteria. We then compared patients with optimal outcomes and those without optimal outcomes (compliant and sufficiently treated vs. non-compliant and insufficiently treated). Continuous variables were compared using Student’s unpaired t test. Categorical variables were compared using the χ^2^ test. Multivariate logistic regression analysis was conducted to assess the impact of relevant covariates on the observed differences in treatment outcomes with APAP. Statistical tests were performed using GNU PSPP 1.4.1 Free Software Foundation, Boston, MA. A p-value of ≤0.05, two tailed was considered statistically significant.

## Results

Table 1 shows the baseline characteristics of the 172 study participants. They were majority male, middle to older aged, obese and generally had severe OSA; most were reported to have loud disruptive snoring. The majority of the cohort had comorbid hypertension, 30.2% had a history of depression and 25.0% had comorbid diabetes mellitus.

**Table 1.** Baseline Characteristics of study participants

|  |  |
| --- | --- |
|  | n = 172 |
| Age (years) | 63.4 ± 13.4 |
| Gender (male) (%) | 54.0 |
| Race (non-Hispanic white) (%) | 65.1 |
| BMI (kg/m<sup>2</sup>) | 35.7 ± 8.6 |
| Hypertension (%) | 63.4 |
| Congestive Heart Failure (%) | 7.6 |
| Atrial Fibrillation (%) | 9.3 |
| Cerebrovascular Accident (%) | 5.8 |
| Diabetes Mellitus (%) | 25.0 |
| Coronary Artery Disease (%) | 14.0 |
| Depression (%) | 30.2 |
| Regular BZD use (%) | 7.6 |
| Baseline AHI (events/hour) | 51.4 ± 30.5 |
| Snoring severity (severe)(%) | 62.4 |
| Apnea Index (events/hour) | 20.9 ± 26.7 |
| Obs Apnea Index (events/hour) | 18.4 ± 24.8 |
| Hypopnea Index (events/hour) | 31.1 ± 18.8 |
| Central Apnea Index (events/hour) | 1.7 ± 3.9 |
| Oxygen nadir (%) | 78.6 ± 9.1 |

Treatment effectiveness and compliance data are displayed in Table 2. Overall, 37.8% of patients had residual AHI ≥ 5 on APAP therapy, with 40.2% of participants being non-compliant. They also had average mask leak durations of 6.7 ± 27.3 minutes of large leaks per hour suggesting that patients spent 11% to 45% of their time on APAP with large leaks.

**Table 2.** APAP Effectiveness and Compliance Data

|  |  |
| --- | --- |
| Residual AHI on PAP (events/hour) | 5.4 ± 4.42 |
| Residual AHI ≥ 5 on PAP (n, %) | 65, 37.8 |
| Mask Type (nasal)(%) | 57.4% |
| Leak Duration (min/day) | 13.9 ± 27.7 |
| Leak Duration/Total nightly usage (min/hr) | 6.7 ± 27.3 |
| Leak Duration/90th percentile pressure (min/cmH2O) | 0.7 ± 2.7 |
| Percentage 4-hour usage per night | 69.1 ± 30.5 |
| <70% 4-hour usage/night (n, %) | 69, 40.2 |
| Mean Pressure (cmH2O) | 8.8 ± 2.3 |
| Peak Avg Pressure (cmH2O) | 10.7 ± 2.6 |
| 90<sup>th</sup> Percentile Pressure (cmH2O) | 10.6 ± 2.5 |
| Clinic Follow-up period (mon) | 5.5 ± 3.6 |

Table 3 shows a comparison of clinical and anthropometric data as a function of whether APAP treatment was effective. The patients with insufficiently treated OSA on APAP (Residual AHI (RAHI) ≥ 5) were older (68.4 ± 12.5 vs. 60.4 ± 13.1 years, p < 0.01) and had lower body mass indices (31.9 ± 6.3 vs. 37.9 ± 9.1 kg/m^2^ , p < 0.01). They had slightly more central apneas at baseline (2.5 ± 4.2 vs 1.2 ± 3.6 events per hour, p=0.046), modestly higher peak PAP pressures (11.4 ± 2.6 vs 10.3 ± 2.4, p = 0.013) and had much longer durations of mask leaks per hour of usage (14.1 ± 42.7 vs 2.1 ± 6.2 minutes per hour, p < 0.01). However, we did not find an association between mask types and higher residual AHI. There was however >50% missing data on mask types. They were also less compliant with APAP when comparing their 4-hour nightly usage (61.2 ± 33.6 vs 73.9 ± 27.4%, p < 0.01). Following multivariate analyses, BMI (OR: 0.85, 95% CI: 0.77-0.93, p <0.01), baseline central apnea index (OR: 1.11, 95% CI: 1.01-1.21, p = 0.04), leak duration (OR: 1.04, 95% CI: 1.02-1.06, p <0.01), APAP compliance (OR: 0.98, 95% CI:0.96 -0.99, p=0.048) and peak average pressure (OR: 1.29, 95% CI: 1.11-1.46, p<0.01) were associated with insufficiently treated OSA on APAP.

**Table 3.** Comparison of groups based on achieving effective AHI (< 5 /h) on APAP

|  | RAHI < 5 | RAHI ≥ 5 |  |
| --- | --- | --- | --- |
| Age (years) | 60.4 ± 13.1 | 68.4 ± 12.5 | <0.01 |
| Gender (% men) | 48.6 | 63.1 | 0.06 |
| BMI (kg/m<sup>2</sup>) | 37.9 ± 9.1 | 31.9 ± 6.3 | <0.01 |
| HTN | 58.9 | 70.8 | 0.11 |
| DM | 29 | 18.5 | 0.12 |
| Depression | 29.9 | 30.8 | 0.90 |
| BAHI (events/hr) | 52.1 ± 32.2 | 50.3 ± 27.6 | 0.71 |
| Snoring Severity n = 133 (severe) | 65.1 | 39.4 | 0.44 |
| CAI (events/hr) | 1.2 ± 3.6 | 2.5 ± 4.2 | 0.046 |
| AI | 19.7 ± 27.4 | 22.8 ± 25.8 | 0.47 |
| AI/HI | 1.8 ± 6.9 | 1.6 ± 3.5 | 0.87 |
| Mask Type n = 68 (nasal) | 61 | 51.9 | 0.46 |
| Leak Duration (min/day) n = 162 | 6.6 ± 13.8 | 25.9 ± 38.8 | <0.01 |
| Leak Duration/total nightly usage (min/hour) n = 160 | 2.1± 6.2 | 14.1± 42.7 | <0.01 |
| Leak Duration /90th percentile pressure (min/cmH2O) n = 160 | 0.2 ± 0.5 | 1.4 ± 4.3 | <0.01 |
| 4 hour usage (%) n = 164 | 73.9 ± 27.4 | 61.2 ± 33.6 | <0.01 |
| Mean Pressure (cmH2O) n = 164 | 8.6 ± 2.3 | 9.1 ± 2.4 | 0.23 |
| Peak Avg Pressure<br>(cmH2O) n=161 | 10.3 ± 2.4 | 11.4 ± 2.6 | 0.013 |
| 90<sup>th</sup> Percentile P<br>(cmH2O) n = 162 | 10.3 ± 2.4 | 11 ± 2.6 | 0.07 |

As shown in Table 4, the non-compliant group of patients were younger and comprised mostly females. There were few anthropometric and clinical differences between non-compliant and compliant patients. However, non-compliance was associated with longer leak durations (13.9 ± 42.1 vs. 1.8 ± 3.5 minutes per hour of usage, p<0.01). Although a higher percentage of non-compliant patients had comorbid depression, the difference only approached statistical significance. Furthermore, the presence of common cardiovascular comorbidities such as hypertension and diabetes mellitus was not associated with differences in compliance in our study. However, in multivariate modelling, only age (OR: 0.96, 95% CI:0.94-0.98, p <0.01) and possibly female gender (OR: 0.51, 95%CI -0.19-1.22, p= 0.058) were associated with APAP compliance.

**Table 4.**
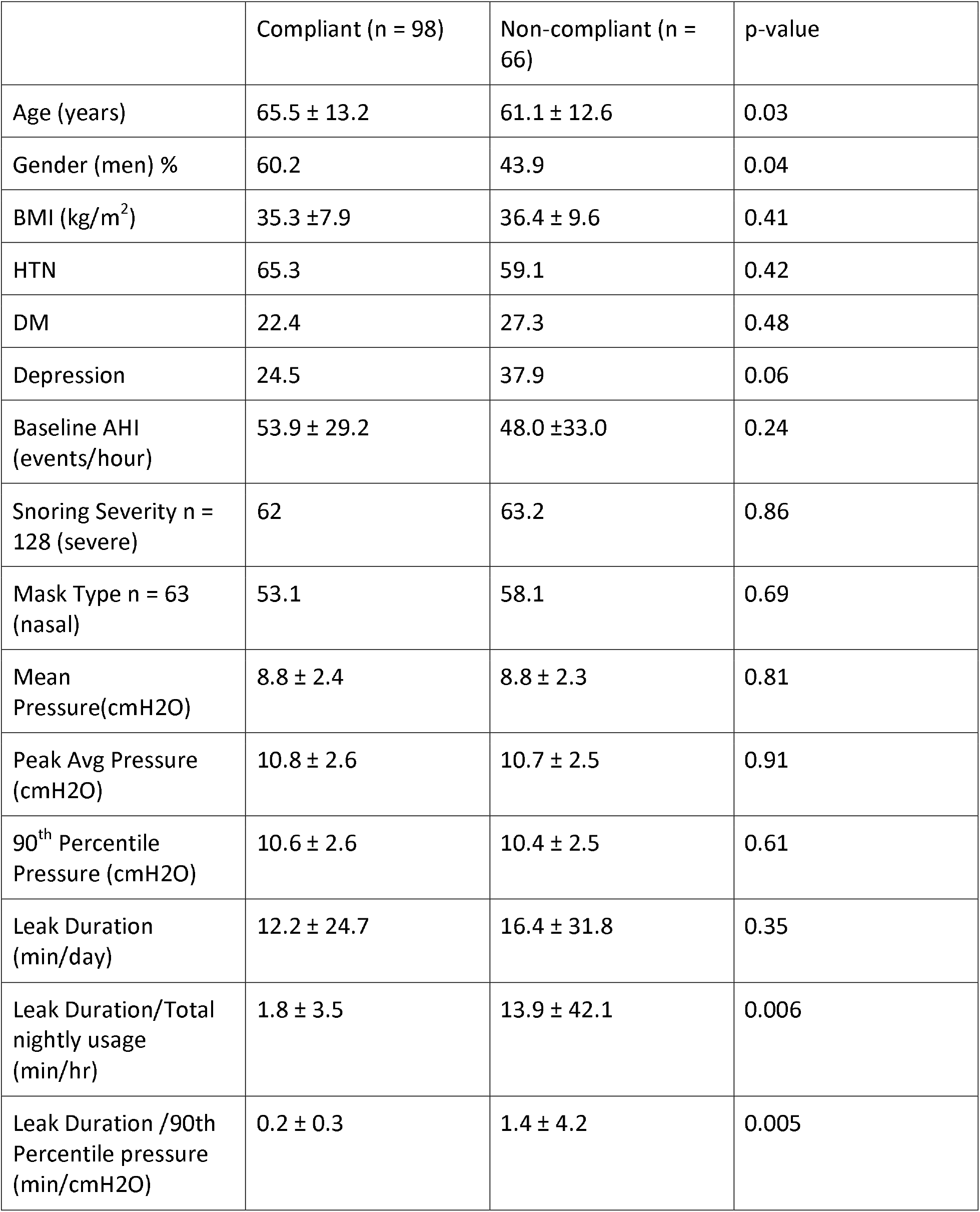
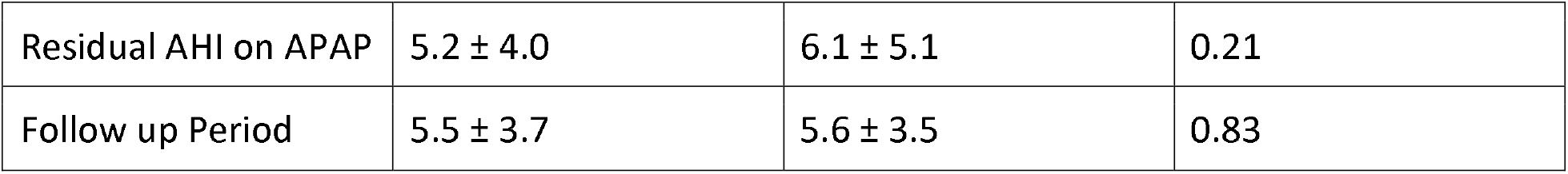
Comparison of groups based on APAP Treatment Compliance

|  | Compliant (n = 98) | Non-compliant (n = 66) | p-value |
| --- | --- | --- | --- |
| Age (years) | 65.5 ± 13.2 | 61.1 ± 12.6 | 0.03 |
| Gender (men) % | 60.2 | 43.9 | 0.04 |
| BMI (kg/m<sup>2</sup>) | 35.3 ± 7.9 | 36.4 ± 9.6 | 0.41 |
| HTN | 65.3 | 59.1 | 0.42 |
| DM | 22.4 | 27.3 | 0.48 |
| Depression | 24.5 | 37.9 | 0.06 |
| Baseline AHI (events/hour) | 53.9 ± 29.2 | 48.0 ± 33.0 | 0.24 |
| Snoring Severity n = 128 (severe) | 62 | 63.2 | 0.86 |
| Mask Type n = 63 (nasal) | 53.1 | 58.1 | 0.69 |
| Mean Pressure (cmH<sub>2</sub>O) | 8.8 ± 2.4 | 8.8 ± 2.3 | 0.81 |
| Peak Avg Pressure (cmH<sub>2</sub>O) | 10.8 ± 2.6 | 10.7 ± 2.5 | 0.91 |
| 90<sup>th</sup> Percentile Pressure (cmH<sub>2</sub>O) | 10.6 ± 2.6 | 10.4 ± 2.5 | 0.61 |
| Leak Duration (min/day) | 12.2 ± 24.7 | 16.4 ± 31.8 | 0.35 |
| Leak Duration/Total nightly usage (min/hr) | 1.8 ± 3.5 | 13.9 ± 42.1 | 0.006 |
| Leak Duration /90th Percentile pressure (min/cmH<sub>2</sub>O) | 0.2 ± 0.3 | 1.4 ± 4.2 | 0.005 |
| Residual AHI on APAP | 5.2 ± 4.0 | 6.1 ± 5.1 | 0.21 |
| Follow up Period | 5.5 ± 3.7 | 5.6 ± 3.5 | 0.83 |

Table 5 shows a comparison of patients who were optimally treated (compliant and adequately treated with APAP) and those who were not optimally treated (non-adherent and insufficiently treated with APAP). The non-optimally treated group had lower BMI (33.2 ± 7.8 vs 37.5 ± 8.3 kg/m2, p = 0.02), higher leak duration per hour of usage (26.9 ± 60.8 vs 1.1 ± 2.2 minutes per hour, p < 0.01) and lower nightly usage (2.5 ± 1.4 vs 6.9 ± 1.7 hours, p < 0.01).

**Table 5.** Comparison of Groups based on Outcomes on APAP

|  | Optimally treated<br>(n= 65) | Not optimally treated<br>(n=29) | p-value |
| --- | --- | --- | --- |
| BMI | 37.5 ± 8.3 | 33.2 ± 7.8 | 0.02 |
| Leak Duration<br>(min/day) | 7.3 ± 16.5 | 30.7 ± 44.1 | <0.01 |
| Leak Duration/Total<br>nightly usage<br>(min/hr) | 1.1 ± 2.2 | 26.9 ± 60.8 | <0.01 |
| Leak Duration /90th<br>Percentile pressure<br>(min/cmH2O) | 0.1 ± 0.2 | 2.75 ± 6.1 | <0.01 |
| Nightly usage (hr) | 6.9 ± 1.7 | 2.5 ± 1.4 | <0.01 |

## Discussion

To our knowledge, this is the first study that has examined the two major factors predicting treatment outcomes (AHI reduction and compliance) in a cohort of patients exclusively treated with APAP. We found that several clinical, demographic and anthropometric factors were associated with worse treatment effectiveness and compliance and that large air leaks were of particular importance.

Our study showed that 37.8% of study participants were insufficiently treated with APAP, evidenced by significant residual sleep apnea (AHI ≥ 5 per hour). This is somewhat higher than has been reported in prior similar studies. One study reported 22.1% [3] and another 32.2% [8]. These differences in findings may be attributable to variations in patient demographics and study methods. For instance, the aforementioned studies had a younger population with average ages of 58.2 and 54.7 years respectively compared to our study which had an average age of 63.4 years. This is particularly relevant as one study examining data from the French national sleep database [3] demonstrated an association between older age and insufficiently treated OSA which is consistent with our findings. However, the French study excluded patients with mild OSA in contrast to our study population which included the entire spectrum of OSA severity. The second study excluded patients with high leak rates on their compliance data. Similar to our study, both studies used device generated AHI, however their study populations were not specifically limited to patients using the auto-adjusting CPAP delivery mode.

The insufficiently treated group also had lower body mass indices compared to the group with optimal AHI. The influence of age and BMI in determining responsiveness to APAP may be indicative of the previously reported heterogeneity of OSA [9] and may play an important role in targeted OSA therapy. Older patients and patients with lower BMIs may have OSA endotypes that may not respond as well to the current APAP algorithms and likely need closer follow up after initiation of APAP or may benefit from alternative modes of PAP delivery or APAP algorithms. For instance, detection of vibrations from snoring is one of the signals utilized in some APAP based algorithms [10] and patients with lower BMI typically snore less. Therefore, one possibility is that the APAP algorithms may be less likely to detect respiratory events associated with less snoring. Furthermore, younger age, female sex and higher BMI have been associated with shorter duration of sleep disordered breathing events [11]. It is therefore possible that older patients and those with lower BMI need a more sustained increase in positive airway pressure delivery during respiratory events than that which is currently provided with current APAP algorithms. Older patients have been described as having an OSA endotype in which upper airway collapsibility plays a bigger role [12] and the variation in pressure delivery by APAP may leave more room for upper airway collapse when compared to the fixed pressure mode in these patients.

Polysomnographic data from the insufficiently treated group also revealed slightly higher central apnea indices on their baseline studies. Although this observation is consistent with prior findings [8] [13], the difference we observed in central apnea indices is unlikely to be clinically relevant or useful.

Analysis of APAP compliance data in our patients with insufficiently treated OSA revealed that they had higher peak average pressure requirements and longer mask leak durations, even after adjusting leak time for total nightly usage and 90th percentile pressure. Higher peak pressures have not typically been associated with higher residual AHIs in prior studies although one study reported that patients with CPAP pressure requirements of > 10 cm [3] had a higher residual AHI.

We found that greater air leak was associated with a higher likelihood of ineffective treatment. Prior data examining the contribution of leaks to higher residual AHI were attributed to the use of oronasal masks. These studies found oronasal masks were associated with higher leaks when compared to nasal masks [14] and nasal pillows [15]. While we found an association between higher residual AHI on APAP with increased leak, we did not find an association with mask type.

Similar to findings in prior studies [3] [13], we also found that the insufficiently treated group, not surprisingly, were less compliant with PAP therapy. One possibility is that low nightly usage hours can skew the AHI calculation due to small denominator values, similar to what is observed with split night studies [16]. Another possibility is that the presumed lack of clinical resolution of OSA symptoms related to a significant residual AHI precluded continued patient compliance with APAP. For instance, data from the SAVE trial found improvement in sleepiness, which is a common symptom of OSA, to be a predictor of compliance with CPAP therapy [17].

We found that 40.2% of our cohort were non-compliant. This is in accordance with prior studies which utilized the same compliance criteria and reported non-compliance rates of 37% [18] and 35.5% [19] respectively. Generally, reported CPAP non-adherence rates have been highly variable, ranging from 29 to 83% [20]. Data on factors predicting adherence are also inconsistent [21]. While there were very few demographic, anthropometric or clinical differences between non-compliant and compliant patients, we found that younger patients and females were less likely to be compliant. It has been well documented that younger age is associated with worse adherence [22] and increasing age with improved adherence [23] [24]. One prior study reported that older males with more severe OSA were more likely to be compliant [25]. While several studies have reproduced the association between CPAP adherence and OSA severity [26], we did not observe increased adherence to therapy with increasing severity of OSA. This may have been because of the preponderance of severe OSA (>70%) in our cohort.

Non-compliance was also associated with longer leak durations. This finding has been documented in a prior study [27]. Another study documented poor adherence to APAP associated with higher leak volume adjusted for pressure delivery [28]. However prior data exploring the effects of mask leaks on CPAP compliance have been inconsistent. One study looking at the effects of types of CPAP interface on adherence documented that even though oronasal masks were associated with higher mask leaks, the adherence rates between different mask interfaces were not significantly different [14]. Another study observed that mask related side effects, of which mask leak was a common complaint, had a negative impact on adherence with PAP [22]. Clinical experience suggests that mask leaks may be disruptive to both patients’ and bedpartners’ sleep thus contributing to PAP intolerance and partner/spousal discord. Spousal involvement has been shown to play a positive role in CPAP compliance [29] and hence spousal discord would be counterproductive. Furthermore, CPAP leaks are more likely to occur at higher CPAP pressure levels [30] which may lead to pressure discomfort and result in poor CPAP tolerance. Nevertheless, we did not find an association between CPAP pressure requirements and compliance.

We acknowledge that a major limitation of our study was that it was retrospective, non-randomized and we had > 50% missing data on mask types which may have resulted in a type 2 error. Despite the aforementioned limitations, our study examined important predictors of OSA treatment outcomes in a large cohort of patients using the increasingly popular auto-adjusting mode of CPAP delivery.

In conclusion, we found several demographic, anthropometric and clinical characteristics associated with worse APAP effectiveness and compliance. These observations have potential implications in the care of patients prescribed APAP. Patients with lower BMI and central apneas at baseline may benefit from early monitoring of their PAP data and treatment adjustments performed either remotely or at early follow up visits to ensure adequate control of OSA. Younger patients may require closer follow up as well. Early targeted behavioral and clinical interventions to improve compliance may be indicated particularly in this group of patients who are at risk of long-term cardiovascular consequences of untreated OSA. Finally, the detrimental effects of large mask leaks appear to be a commonality impacting both treatment effectiveness and adherence and thus affecting APAP treatment outcomes. Timely and ongoing interventions to minimize mask leaks may thus have a vital role to play in improving APAP treatment outcomes in OSA. Clinical interventions which focus on careful mask fitting and enhanced training practices on proper mask application techniques during PAP delivery will likely be beneficial. Creating systems to flag and track patients with subjective leak complaints and/or significant leaks on PAP data for early follow up and re-fitting by respiratory technologists may help to address leaks early enough to reduce APAP therapy dropout rates.

## Data Availability

All data produced in the present study are available upon reasonable request to the authors

## Abbreviations

CPAP: Continuous Positive Airway Pressure
APAP: Auto-titrating Positive Airway Pressure
OSA: Obstructive Sleep Apnea
AHI: Apnea Hypopnea Index
RAHI: Residual AHI
BAHI: Baseline AHI

